# Subthreshold Nanosecond Laser Treatment of Intermediate AMD without Reticular Pseudodrusen: Real-world Three-year Follow-up Study

**DOI:** 10.1101/2022.05.29.22275655

**Authors:** David R. Worsley, Jennie Louise, Susan P. Bull

**Affiliations:** Hamilton Eye Clinic, Hamilton, New Zealand; Faculty of Health Sciences, University of Adelaide, Australia

## Abstract

**Purpose:** Subthreshold nanosecond laser (SNL) has been proposed to reduce the risk of intermediate age-related macular degeneration (iAMD) progressing to late AMD. The phase 3 LEAD Study post-hoc analysis indicates a potentially large benefit from SNL treatment of eyes without reticular pseudodrusen. This real-world study reports the three-year outcomes of SNL treatment of iAMD without RPD.

**Methods:** An observational retrospective single-centre cohort study of all patients with iAMD, centre-involving very large soft drusen (≥250 μm diameter) without RPD, treated with SNL and with three-year follow up. The primary outcome measured was progression to late AMD; neovascular AMD (nAMD) or geographic atrophy (GA).

**Results:** There were 120 eyes of 64 patients. At baseline, the cohort had a high risk profile; drusen median area, volume and largest diameter were 0.70mm^2^ (IQR: 0.20 to 1.50), 0.03mm^3^ (IQR: 0.01 to 0.08) and 835μm (IQR: 446.50 to 1398.50) respectively; hyperreflective foci were present in 56.67%; and hyporeflective drusen cores in 25.83%. Eyes had a mean of 3.03 treatments. By three years, progression to late AMD occurred in 5.83% of eyes, all to GA. Visual acuity was stable or improved in 80% of eyes.

**Conclusion:** The three-year progression rate was low compared with published 36-month natural histories of iAMD without RPD. The progression rate was similar to the LEAD study SNL-treated group of iAMD without RPD. This study supports the hypothesis that SNL for iAMD without RPD may reduce progression to late AMD. Further investigation is warranted.

## Introduction

Intravitreal anti-VEGF therapy has led to a major reduction in vision loss from neovascular AMD (nAMD), however there is no proven therapy for geographic atrophy (GA).^1^ Altering modifiable lifestyle risk factors are associated with slowing of the earlier disease stages. AREDS-2 supplementation has a modest effect on slowing progression to late-stage nAMD, but not to GA.^2^ There remains an unmet need for effective therapies to reduce the risk of progression from early to late-stage AMD.

The hallmark of early AMD is progressive focal accumulation of abnormal extracellular debris between the retinal pigment epithelium (RPE) and Bruch’s membrane (BM), clinically apparent as soft drusen.^3^ The classification of AMD is: early AMD with medium soft drusen (63-125 μm diameter) and no pigment abnormalities; intermediate AMD (iAMD) with large soft drusen (≥125 μm diameter) or medium soft drusen with pigment abnormalities; and late AMD with nAMD or GA.^4^

In early and iAMD, the drusen load (number, area and volume) progressively increases over time.^5^ Soft drusen load is a major biomarker for risk of progression to late AMD.^6^ At the high end of the risk spectrum are extremely large soft drusen, termed drusenoid pigment epithelial detachment (DPED).^7^ Additionally, pigmentary changes, hyperreflective foci (HRF) and hyporeflective drusen cores (HDC) are high-risk SD-OCT biomarkers.^6, 8, 9^

Although not included in the classification, reticular pseudodrusen (RPD), focal accumulations of debris in the subretinal space, is now recognized as a phenotype with a high risk of progression.^8, 10^ Furthermore, RPD has been proposed as a separate disease pathway to late AMD.^11^

Following observations that continuous wave (CW) laser leads to soft drusen regression, studies looked at whether this reduces the risk of progression to late AMD. A Cochrane review of pooled data from 11 RCTs concluded that, although CW laser leads to drusen regression, it does not influence progression to late disease, nor result in an increased risk of nAMD, GA or vision loss.^12^ On the downside, CW laser causes RPE and outer retinal destruction with scar formation, which may lead to choroidal neovascularization.

Subthreshold laser is delivery of a very short laser pulse duration to substantially reduce energy delivery, sufficient for RPE cell damage but without collateral damage to adjacent tissues.^13^ Below a 4 ms pulse threshold, thermal damage is believed to be entirely intracellular with no collateral damage together with non-thermal mechanical damage from small bubbles of steam formation adjacent to melanosomes.^14^ The therapeutic effect is considered to arise from RPE cell repair processes.

2RT (AlphaRET, Adelaide, Australia) is a Q-switched, frequency doubled laser delivering a 3 ns pulse.^15^ A 400-μm diameter speckled beam profile results in variable fluence within the laser spot. Animal (in-vivo and ex-vivo) and human eye (ex-vivo) studies showed that, at clinically relevant doses, RPE cell apoptosis within the treatment spot was both sporadic and selective, and without collateral damage to adjacent tissues.^16-19^ RPE regeneration was by dedifferentiation, proliferation, and migration of surrounding RPE cells.

In a pilot study, a single SNL treatment of patients with iAMD was associated with a reduction in drusen load, without evidence of progression and without clinical evidence of photoreceptor damage.^20^ The phase 3 LEAD study compared 6-monthly SNL treatment to sham treatment in 292 patients with iAMD and large soft drusen.^21, 22^ At 36 months follow-up there was no overall delay in the rate of progression to late AMD. A post-hoc effect modification analysis indicated that SNL was highly effective in the RPD-phenotype, with a >4-fold reduction in progression to late AMD. As the post-hoc analysis is biologically plausible, the authors concluded that SNL may slow progression of the RPD-phenotype, however this would need further validation. An observational 24-month extension study showed that the RPD-phenotype had a persistence of the potential benefit.^23^ With evidence indicating a potential positive benefit and excellent safety, the FDA has recently provided guidance on the registration pathway for SNL treatment of the RPD-phenotype.^24^

Here we report a single-centre, retrospective, three-year follow-up consecutive case series of 120 eyes with iAMD, large soft drusen and RPD-phenotype, treated with SNL.

## Methods

A retrospective observational single-centre study was undertaken at Hamilton Eye Clinic, a tertiary referral centre in New Zealand. Written study consent was obtained for all patients, except for those lost to follow-up. As the study uses only de-identified data with no active human participants, review was waived by the Health and Disability Ethics Committee. The study and clinical care complied with the Declaration of Helsinki.

### Participants

All participants were identified from the electronic records at Hamilton Eye Clinic. Assessment of baseline imaging immediately prior to the first SNL treatment was used to assess eligibility. For this purpose, spectral-domain OCT (SD-OCT) imaging (Spectralis, Heidelberg Technologies; high resolution volume scan, 15°x 15°, ART 15 frames and 37 sections) and fundus autofluorescence (FAF) were analyzed.

Included were patients with very large soft drusen (≥250 μm diameter) present within a 3000 μm diameter circle centered on the fovea. Excluded were patients with the RPD+ phenotype, defined as: in either eye, >5 definite SD-OCT RPD on a single OCT slice or definite RPD on FAF.^10,21^ Excluded were patients with clinical features, in either eye, of nAMD or GA (as defined by Sadda et al^25^). Nascent GA (as defined by Wu et al^26^) was not excluded. Also excluded were eyes with current ocular disease or past treatment that may influence the natural history of AMD.

To reduce positive bias, eyes were selected for inclusion before the clinical file was accessed for last follow-up and outcome data.

### Baseline data

Data collected at baseline included gender, age, family history, smoking history, AREDS supplementation, and Snellen visual acuity (converted to logMAR for statistical analysis).

For each eye, important prognostic features present within the central 3000 μm diameter circle on SD-OCT imaging were recorded: largest drusen diameter, drusen area and volume (calculated using ‘Advanced RPE Analysis’ of the Cirrus SD-OCT 512 × 128 cube 6×6 mm), and presence of HRF or HDC.

### SNL treatment

Laser fluence was set at the lower of 70% of threshold or 0.24 mJ. Following the positive LEAD study interim safety report the number of spots was increased from 12 to 50.^27^ Spots were placed in a double row temporal arc outside a 4000μm diameter circle centered on the fovea. All laser spots were spaced by greater than a two-spot diameter.

### Follow-up and SNL re-treatment

Patients with higher-risk features were recommended a 6-month follow-up, while those with lower-risk features were recommended an annual follow-up. Snellen visual acuity was recorded. Any progression to late disease was assessed using SD-OCT: nAMD by typical SD-OCT features, such as intraretinal fluid, subretinal fluid or hemorrhage; GA by SD-OCT features of complete RPE and outer retinal atrophy (cRORA).^25^

As proposed by Garcia-Filho et al^28^, changes in drusen load were used as a biomarker of presence or absence of a treatment effect. Based on this, re-treatment was offered for eyes with increasing or unchanged drusen load, and not offered for eyes with drusen regression.

Re-treatment laser parameters were adjusted according to laser-induced RPE changes seen on FAF. Treatment power was modestly increased if there were no changes, unaltered if there was hypofluorescence and reduced if there was hyperfluorescence.

### Last follow-up data

Last visit was either at three years (34-36 months), or when progression was observed or lost to follow-up. Data collected at last visit was Snellen visual acuity, months of follow-up, drusen area and volume, any progression to late disease, and number of treatments.

### Statistical analysis

Statistical analysis was performed using Stata v16 (StataCorp, College Station, TX). Baseline characteristics of patients were assessed descriptively. Continuous variables were summarized using means and standard deviations, or medians and interquartile ranges, as appropriate. Categorical variables were summarized as frequencies and percentages. The risk of progression was modelled using log binomial regression, with Generalized Estimating Equations to account for correlation due to patients with two eyes included in the study. An offset was also included to account for variable length of follow-up time. The estimates from these models are the overall risk of progression within three years, presented as a percentage, and 95% Confidence Interval. The strength of association between potential prognostic factors (drusen diameter ≥1000*μ*m, hyperreflective foci and hyporeflective drusen cores) was assessed descriptively and using Fisher’s Exact test due to the small number of progressions.

## Results

One hundred and twenty eyes in 64 patients were identified. The baseline patient and eye characteristics are in Table 1. The mean and median months of follow-up were 36.00 months (IQR 36.00, 36.00) and 36.13 (SD 7.89) respectively. The mean number of treatments was 3.03 (range 1-5, SD 1.03). For baseline largest drusen diameter of 250-349μm, 350-999μm and ≥1000μm, the mean number (SD) of treatments was 2.52 (0.81), 2.93 (0.98) and 3.32 (1.07) respectively.

**Table 1:** Patients Baseline Characteristics.

| Patients |  |
| --- | --- |
| Number | 64 |
| Female: N(%) | 40 (62.50) |
| Family History: N(%) | 28 (43.75) |
| Smoker: N(%) | 9 (14.06) |
| AREDS: N(%) | 17 (26.98) |
| Bilateral SNL treatment: N(%) | 59 (92.19) |
| Eyes |  |
| Number | 120 |
| Age: Mean (SD) | 68.73 (9.02) |
| logMAR: Mean (SD) | 0.11(0.12) |
| Drusen Area (square root) mm (central 3mm): Median (IQR) | 0.70 (0.20, 1.50) |
| Drusen Volume (cube root) mm (central 3mm): Median (IQR) | 0.03 (0.01, 0.08) |
| Largest Drusen Diameter: |  |
| All eyes: N, Median μm (IQR) | 120, 835.00 μm (446.50, 1398.50) |
| 250-299μm: N, %, Median μm (IQR) | 21, 17.5, 304.00 μm (261, 326) |
| 350-999μm: N, %, Median μm (IQR) | 46, 38.33, 602.50 μm (467, 761) |
| ≥1000μm: N, %, Median μm (IQR) | 53, 44.17, 1480.00 μm (1183, 1640) |
| Hyperreflective Foci: N (%) | 68 (56.67) |
| Hyporefective Drusen Cores: N (%) | 31 (25.83) |

### Progression to late AMD

Using raw data, progression to late AMD within three-years was seen in seven eyes of seven patients (5.83% of eyes; 95% confidence interval: 2.76, 11.81), all to GA and none to nAMD. No patient with bilateral treatment had bilateral progression.

The estimated risk of eye progression is 5.82% (95% CI: 2.55, 13.30) with the log binomial regression model with GEEs and offset for variable follow-up time. With the Adjusted GEE + Offset model the estimated risk is 5.10% (95% CI: 0.37, 9.33). The cumulative hazard function is shown in Figure 1. As fewer than 50% of eyes progressed, estimating median time to progression is not possible. Largest drusen diameter, HRF and HDC were each strongly associated with risk of progression at three years (Table 2). All seven eyes that progressed had a baseline drusen diameter ≥1000μm. The interrelationship of largest drusen diameter with other measures of drusen size (drusen area and volume), with other risk factors for progression (HRF and HDC), and with visual acuity outcomes is shown in Table 3.

**Table 2:**
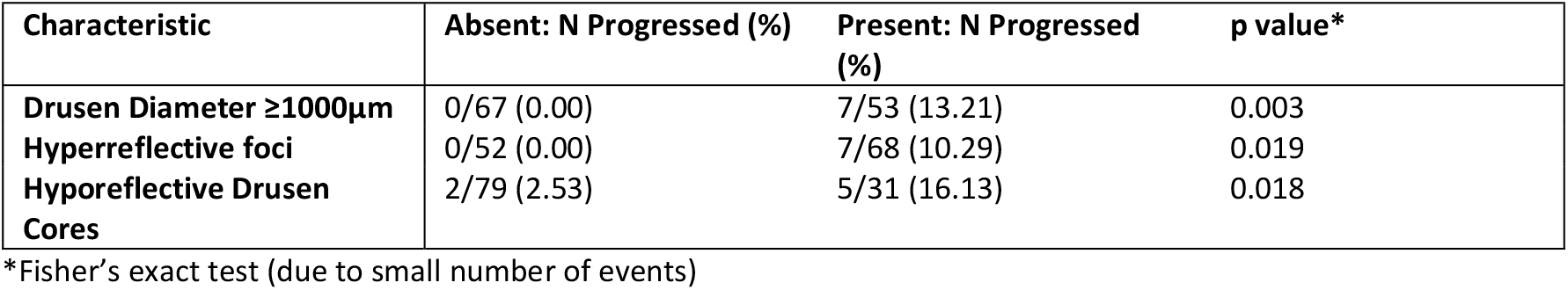
Eye Baseline Characteristics: Association with Progression to Late AMD.

| Characteristic | Absent: N Progressed (%) | Present: N Progressed (%) | p value* |
| --- | --- | --- | --- |
| Drusen Diameter $\geq 1000\mu\text{m}$ | 0/67 (0.00) | 7/53 (13.21) | 0.003 |
| Hyperreflective foci | 0/52 (0.00) | 7/68 (10.29) | 0.019 |
| Hyporefective Drusen Cores | 2/79 (2.53) | 5/31 (16.13) | 0.018 |

\*Fisher's exact test (due to small number of events)

**Table 3:** Eye Baseline Characteristics: Drusen Diameter.

| | Diameter<br>250 - 349$\mu\text{m}$ | Diameter<br>350 - 999$\mu\text{m}$ | Diameter<br>$\geq 1000\mu\text{m}$ | Overall |
| --- | --- | --- | --- | --- |
| n (Eyes) | 21 (17.50) | 46 (38.33) | 53 (44.17) | 120 |
| Drusen Diameter: Median (IQR) | 304.00 (261.00, 326.00) | 602.50 (467.00, 761.00) | 1480.00 (1183.00, 1640.00) | 835.00 (446.50, 1398.50) |
| Drusen Area $\text{mm}^2$ (central 3mm): Median (IQR)* | 0.10 (0.00, 0.20) | 0.40 (0.20, 0.70) | 1.45 (1.00, 2.10) | 0.70 (0.20, 1.50) |
| Drusen Volume $\text{mm}^3$ (central 3mm): Median (IQR)* | 0.00 (0.00, 0.01) | 0.01 (0.01, 0.03) | 0.08 (0.04, 0.13) | 0.03 (0.01, 0.08) |
| Hyperreflective Foci: N (%) | 4 (19.05) | 22 (47.83) | 42 (79.25) | 68 (56.67) |
| Hyporefective Drusen Cores: N (%) | 0 (0.00) | 5 (10.87) | 26 (49.1) | 31 (25.83) |
| logMAR: Mean (SD) | 0(0.10) | 0.10 (0.12) | 0.14 (0.12) | 0.11 (0.12) |

\*Drusen area/volume data was available for 103/113 eyes that had not progressed by three years.

**Figure 1.**
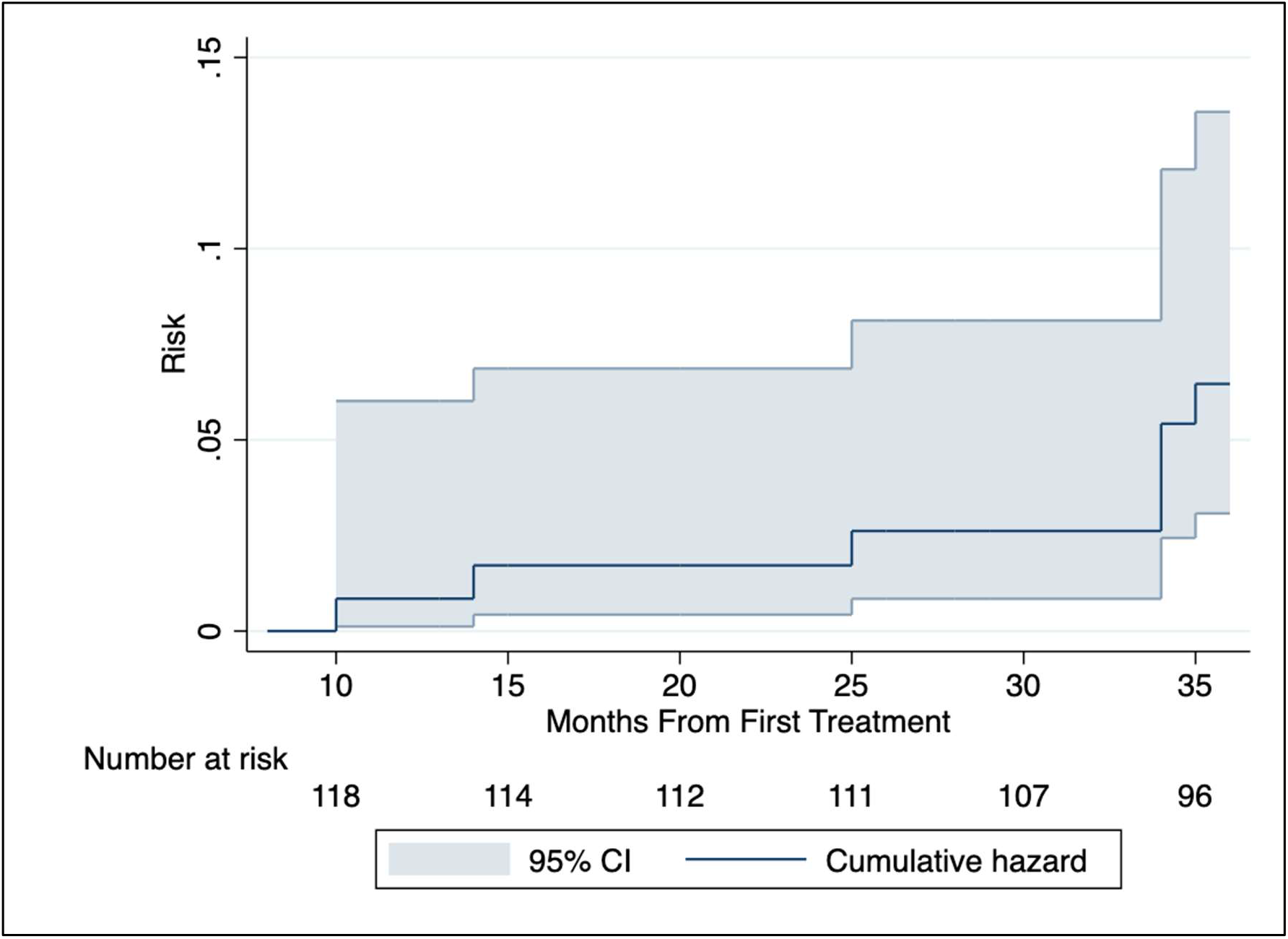
Cumulative Hazard Function.

By three years, baseline drusen area had increased in 71 (68.93%), was unchanged in 6 (5.83%) and decreased in 26 (25.24%) eyes. No eyes with a decrease in drusen area subsequently progressed to late AMD.

### Visual acuity

Visual acuity outcomes are summarized in Table 4. At the final follow-up visit, 96 eyes (80.00%) had either an improvement or no deterioration in visual acuity. Visual acuity declined in 24 eyes (20.00%); 22 eyes (18.33%) by 1-2 lines, and 2 eyes (1.67%) by 3-4 lines. The two eyes (two patients) with 3-4 lines loss had DPED at baseline. By three years both had developed foveal pigmentary abnormality with disrupted outer retinal layers, but without progression to late disease.

**Table 4:** Visual Acuity Results.

| Characteristic | All Eyes (n=120) | Eyes with FU ≥34mths (n=107) * |
| --- | --- | --- |
| **logMAR at baseline** |  |  |
| - Mean (SD) | 0.11 (0.12) | 0.10 (0.12) |
| - Median (IQR) | 0.10 (0.00, 0.20) | 0.08 (0.00, 0.20) |
| **logMAR at last follow-up** |  |  |
| - Mean (SD) | 0.11 (0.12) | 0.09 (0.10) |
| - Median (IQR) | 0.10 (0.02, 0.17) | 0.10 (0.02, 0.14) |
| **Change in logMAR** |  |  |
| - Mean (SD) | 0.01 (0.11) | 0.01 (0.11) |
| - Median (IQR) | 0.00 (-0.06, 0.04) | 0.00 (-0.06, 0.04) |
| **Change in logMAR** |  |  |
| - Gain ≥4 | 1 (0.83) | 0 (0.00) |
| - Gain 3 to <4 | 1 (0.83) | 1 (0.93) |
| - Gain 2 to <3 | 2 (1.67) | 2 (1.87) |
| - Gain 1 to <2 | 15 (12.50) | 15 (14.02) |
| - Gain <1 to Loss <1 | 77 (64.17) | 66 (61.68) |
| - Loss 1 to <2 | 14 (11.67) | 13 (12.15) |
| - Loss 2 to <3 | 8 (6.67) | 8 (7.48) |
| - Loss 3 to <4 | 2 (1.67) | 2 (1.87) |

\*13/120 eyes were either lost to follow-up or progressed at an earlier time

Six patients with ten eyes were lost to follow-up: three deceased and three due to frailty.

No adverse events related to laser application were observed. Two informally documented laser spot-related effects were occasionally documented at follow-up; after-images which resolved over days to weeks, and focal pigment changes at laser spot sites but without overlying outer retinal OCT changes.

## Discussion

This study assessed “real-world” three-year outcomes of patients with iAMD and the RPD-phenotype, treated with SNL.

Progression to late AMD was 5.83% (using the preferred GEE + Offset model), all to GA and none to nAMD. Baseline drusen load, particularly DPED (largest drusen diameter >1000μm), HRF, and HDC, were strongly associated with progression.

The estimated risk of progression, using an intercept-only log binomial regression model, was very close to the observed proportion. This model accounted for correlation due to patients with two eyes included, and for variable length of follow-up time. It was not adjusted for potential confounding factors, as the small number of progressions observed meant that a multivariable model was not recommended. However, an additional sensitivity analysis was carried out in which age, sex, family history, smoking history, AREDS use and drusen area were included as covariates; this model produced results consistent with the main analysis.

The progression rate by three years compares favorably with two previously reported natural history studies with progression rates of iAMD without RPD.^22, 29^ The AREDS Study 2 published Kaplan Meier curves from which estimates of progression to late AMD by three years can be extrapolated to be 18% (nAMD 8%, GA 10%).^29^ The LEAD sham treatment arm had a very similar progression rate of 19.1% (nAMD 3.65%, GA 15.5%).^22^

The LEAD study provides the only comparable published SNL treatment group.^22^ In LEAD, three-year progression was 5.36% (4.5% GA, 0.9% nAMD); this is a very similar rate to this study.

In contrast to both AREDS-2 and LEAD, this study has both a skew toward high-risk features at baseline, and a different threshold for progression to GA. These differences make the comparatively low three-year progression rate in this study even more impressive. The AREDS-2 included all eyes with soft drusen ≥125 μm diameter, while this study is restricted to eyes with very large soft drusen ≥250 μm diameter. In this study, at baseline, 82.53% of eyes have a drusen largest diameter ≥1000μm, 56.67% of eyes have HRF, and 25.83% have HDC. This high-risk skew is due to both a real-world bias for referral of patients with high-risk features, and not offering treatment when iAMD was at the lower end of the risk spectrum. In this study, the definition of progression to GA is any atrophy within the 15°x 15°OCT scan area; this is a lower threshold than the AREDS-2 definition (atrophy involving the fovea).^29^ This study includes nascent GA at baseline, while the LEAD study excluded this.^21^ Consequently, GA is a progression endpoint in our study, whereas both nascent GA and GA were progression endpoints in the LEAD study.

Currently, there is no evidence-based guidance on the optimal SNL treatment protocol; fluence, number of spots, and retreatment interval. Our treatment protocol differed from the LEAD study. Nearly all patients had bilateral treatment. The treatment intervals were individualized: according to risk profile, and real-world patient’s involvement in treatment and follow-up interval decision-making. Treatment was discontinued once drusen regression was observed. The low rate of progression may indicate that SNL has a wide therapeutic window and may be titratable according to progression risk. Of note, LEAD found no evidence of a dose–response relationship.^30^

Visual acuity improved or was stable in 80% of eyes in this study. As natural history studies have found that high-risk features predict a decline in visual function, these results are encouraging.^31-33^ Of those eyes with a visual acuity decline, in all but two eyes this was a modest 1-2 lines. The two eyes (1.67%) with 3-4 lines loss were developing DPED-related foveal disruption. Visual acuity decline is a common outcome for DPED.^31^

No adverse events were observed. The occurrence of temporary after images and asymptomatic pigment changes at laser spot sites have been reported with other subthreshold laser systems.^34^

Limitations of this study include being retrospective and single-center. The observational cohort study design does not necessarily compromise the importance of the findings. A Cochrane Review concluded a real-world observational study of the size and with the robust results of this study is potentially of comparable value to a randomized study.^35^ As described in the methods, measures were taken to reduce the risk of positive bias both in patient selection and in the recording of outcome data. Using natural history studies for a comparative untreated rate of progression is potentially problematic as there are varying definitions, inclusions, and exclusions. Despite these issues, a near 3-fold reduction in the rate of progression, furthermore in a cohort with a baseline skew to high-risk features, indicates that a beneficial effect has been observed.

This real-world, retrospectively studied, SNL-treated cohort has a 5.83% three-year rate of progression to late AMD. This compares very favourably with rates of 18% and 19.1% in comparable natural history studies. This outcome suggests a potential role for SNL to reduce progression of iAMD without RPD to late disease. Further studies are required to confirm a benefit, give guidance for laser dosimetry, and investigate SNL in other AMD phenotypes.

## Data Availability

All data produced in the present work are contained in the manuscript

